# Coronary Microvascular Function Following Severe Preeclampsia

**DOI:** 10.1101/2024.03.04.24303728

**Authors:** Michael C. Honigberg, Katherine E. Economy, Maria A. Pabón, Xiaowen Wang, Claire Castro, Jenifer M. Brown, Sanjay Divakaran, Brittany N. Weber, Leanne Barrett, Anna Perillo, Anina Y. Sun, Tajmara Antoine, Faranak Farrohi, Brenda Docktor, Emily S. Lau, Doreen DeFaria Yeh, Pradeep Natarajan, Amy A. Sarma, Robert M. Weisbrod, Naomi M. Hamburg, Jennifer E. Ho, Jason D. Roh, Malissa J. Wood, Nandita S. Scott, Marcelo F. Di Carli

**Affiliations:** Cardiology Division, Department of Medicine, Massachusetts General Hospital, Harvard Medical School, Boston, MA; Program in Medical and Population Genetics, Broad Institute of MIT and Harvard, Cambridge, MA; Division of Maternal-Fetal Medicine, Department of Obstetrics and Gynecology, Brigham and Women’s Hospital, Harvard Medical School, Boston, MA; Division of Cardiovascular Medicine, Department of Medicine, Brigham and Women’s Hospital, Harvard Medical School, Boston, MA; Cardiovascular Imaging Program, Departments of Radiology and Medicine, and Division of Nuclear Medicine and Molecular Imaging, Department of Radiology, Brigham and Women’s Hospital, Harvard Medical School, Boston, MA; Whitaker Cardiovascular Institute, Boston University School of Medicine, Boston, MA; Division of Cardiovascular Medicine, Department of Medicine, Beth Israel Deaconess Medical Center, Harvard Medical School, Boston, MA; Lee Health Heart Institute, Fort Myers, FL

**Keywords:** preeclampsia, pregnancy, women’s health, cardiac positron emission tomography, coronary microvascular function

## Abstract

**Background:** Preeclampsia is a pregnancy-specific hypertensive disorder associated with an imbalance in circulating pro- and anti-angiogenic proteins. Preclinical evidence implicates microvascular dysfunction as a potential mediator of preeclampsia-associated cardiovascular risk.

**Methods:** Women with singleton pregnancies complicated by severe antepartum-onset preeclampsia and a comparator group with normotensive deliveries underwent cardiac positron emission tomography (PET) within 4 weeks of delivery. A control group of pre-menopausal, non-postpartum women was also included. Myocardial flow reserve (MFR), myocardial blood flow (MBF), and coronary vascular resistance (CVR) were compared across groups. Soluble fms-like tyrosine kinase receptor-1 (sFlt-1) and placental growth factor (PlGF) were measured at imaging.

**Results:** The primary cohort included 19 women with severe preeclampsia (imaged at a mean 16.0 days postpartum), 5 with normotensive pregnancy (mean 14.4 days postpartum), and 13 non-postpartum female controls. Preeclampsia was associated with lower MFR (*β*=-0.67 [95% CI −1.21 to −0.13]; *P*=0.016), lower stress MBF (*β*=-0.68 [95% CI, −1.07 to −0.29] mL/min/g; *P*=0.001), and higher stress CVR (*β*=+12.4 [95% CI 6.0 to 18.7] mmHg/mL/min/g; *P*=0.001) vs. non-postpartum controls. MFR and CVR after normotensive pregnancy were intermediate between preeclamptic and non-postpartum groups. Following preeclampsia, MFR was positively associated with time following delivery (*P*=0.008). The sFlt-1/PlGF ratio strongly correlated with rest MBF (*r*=0.71; *P*<0.001), independent of hemodynamics.

**Conclusions:** In this exploratory study, we observed reduced coronary microvascular function in the early postpartum period following severe preeclampsia, suggesting that systemic microvascular dysfunction in preeclampsia involves the coronary microcirculation. Further research is needed to establish interventions to mitigate risk of preeclampsia-associated cardiovascular disease.

## Introduction

Preeclampsia is a pregnancy-specific disorder characterized by new-onset or worsening hypertension after 20 weeks’ gestation accompanied by proteinuria or other end-organ dysfunction.^1,2^ Up to 8% of child-bearing individuals in the U.S. experience preeclampsia in 1 or more pregnancies.^3^ Preeclampsia represents a leading cause of maternal and infant morbidity and mortality both in the U.S. and globally, with potential maternal complications including stroke, seizure, kidney injury, pulmonary edema, and peripartum cardiomyopathy (PPCM). In addition, preeclampsia portends heightened long-term maternal risk of atherosclerotic cardiovascular disease and heart failure.^4–9^

The late-stage pathophysiology of preeclampsia is characterized by an excess of circulating anti-angiogenic factors (e.g., soluble fms-like tyrosine kinase receptor 1 [sFlt-1] and soluble endoglin) relative to pro-angiogenic factors (e.g., placental growth factor [PlGF] and vascular endothelial growth factor [VEGF]).^10^ This angiogenic imbalance leads to systemic maternal endothelial dysfunction and vasoconstriction, which in turn causes hypertension, proteinuria, and other clinical manifestations of preeclampsia.^11^ Women who develop preeclampsia have reduced flow-mediated dilation, a marker of peripheral endothelial function, in the antepartum setting, at the time of preeclampsia, and potentially up to several years postpartum.^12^ In addition, severe preeclampsia has been associated with differences in cardiac structure and function in the acute setting by echocardiography, including relatively impaired diastolic function, reduced longitudinal strain, and increased left ventricular (LV) wall thickness compared with findings in normotensive pregnant individuals.^13^ Prior work indicates that higher circulating sFlt-1 levels correlate with lower myocardial performance index (an integrated echocardiographic measure of systolic and diastolic function)^14^ and global longitudinal strain at the time of preeclampsia diagnosis independent of blood pressure.^15^ Furthermore, preeclampsia-associated coronary microvascular dysfunction has been postulated to contribute to later-life cardiovascular disease, including heart failure with preserved ejection fraction (HFpEF).^16^ Whether preeclampsia impacts peripartum coronary microvascular function, however, has not been tested to date.

Myocardial flow reserve (MFR), calculated as the ratio of hyperemic myocardial blood flow (MBF) to rest MBF, represents the integrated effects of epicardial coronary artery disease (CAD) and microvascular vasomotor function; in the absence of obstructive epicardial CAD, impairment in MFR reflects coronary microvascular dysfunction. MFR is reproducible and prognostic, stratifying risk of major adverse cardiovascular events and of HFpEF in midlife and elderly adults.^17,18^ Cardiac positron emission tomography (PET) represents the most extensively validated non-invasive modality for assessment of coronary microvascular function.^17^

Therefore, to determine the acute effects of severe preeclampsia on coronary microvascular function, we prospectively enrolled a cohort of postpartum women and compared PET-derived indices of MFR and MBF with values from pre-menopausal, non-postpartum female controls. We hypothesized that MFR would be reduced after delivery in women with preeclampsia. In addition, we assessed clinical, echocardiographic, and angiogenic biomarker correlates of MFR and MBF among women with severe preeclampsia.

## Methods

### Study Cohorts

This study received human subjects approval from the Mass General Brigham Institutional Review Board, and all participants provided written informed consent. Between December 2020 and October 2022, we prospectively recruited and enrolled women aged ≥18 years delivering at Brigham and Women’s Hospital and Massachusetts General Hospital, two large academic medical centers in Boston, Massachusetts. Recruitment was conducted principally during admission to the antepartum or postpartum obstetrical floors; normotensive control subjects were additionally recruited from routine prenatal visits and via an online research recruitment portal. Women with severe antepartum-onset preeclampsia were identified according to American College of Obstetricians and Gynecologists (ACOG) criteria as those with preeclampsia accompanied by severe hypertension (blood pressure ≥160/110 mmHg) after 20 weeks’ gestation accompanied by proteinuria (300 mg/24 h collection, spot urine protein/creatinine ratio ≥0.3 mg/dL, or urine dipstick with 2+ proteinuria) and/or accompanied by other qualifying severe features (thrombocytopenia, renal insufficiency, liver function abnormalities, pulmonary edema, new-onset headache, or visual symptoms).^19^ Given potential overlap in pathophysiology between preeclampsia and preterm birth,^16^ and in order to isolate any potential effects, if present, specifically attributable to pregnancy, the healthy postpartum comparator group included women who delivered at term (≥37 weeks’ gestation) and who lacked chronic hypertension, gestational hypertension, preeclampsia, or gestational diabetes. Exclusion criteria were body mass index ≥50 kg/m^2^ or history of epicardial CAD or coronary artery dissection, valvular heart disease, or pre-existing cardiomyopathy prior to pregnancy. Prospectively enrolled participants underwent PET imaging, transthoracic echocardiography, and phlebotomy as part of a single study visit performed at Brigham and Women’s Hospital as soon as possible following delivery and no later than one month postpartum. Given prior literature suggesting that normal cardiac remodeling and biomarker risk thresholds associated with adverse outcomes differ between singleton and twin pregnancies,^20,21^ participants (*n*=3) with twin gestations were excluded from the present analysis (**Figure S1**).

Measures of MBF and flow reserve by PET imaging in postpartum participants were compared with those from a control group of healthy, non-diabetic, pre-menopausal, non-postpartum research participants enrolled in a prior study using identical methods for assessing myocardial perfusion with PET.^22^ Exclusion criteria included a history of uncontrolled hypertension, diagnosed cardiac or pulmonary disease, cerebrovascular or peripheral artery disease, or laboratory evidence of kidney or hepatic dysfunction.

### Cardiac Positron Emission Tomography

Postpartum participants underwent measurement of MBF performed on a whole-body PET-computed tomography scanner (Discovery MI, GE Healthcare, Milwaukee, WI) at rest and during vasodilator stress using ^13^N-ammonia or ^82^Rb-rubidium as the flow tracer. Prior work suggests high reproducibility across radiotracers.^23^ The protocol was designed to ensure safety and acceptability for newly postpartum individuals. Adenosine was used as the vasodilator agent to minimize interruption of breastfeeding. Studies were performed after 4 hours of fasting and at least 12 hours following last caffeine intake. Participants were generally advised to withhold antihypertensive medications on the morning of the study visit; however, given the inclusion of women with severe preeclampsia in the early postpartum period (a high-risk period for persistent hypertension and associated complications^24^), we permitted participants to take antihypertensive medications if deemed medically necessary by the study team or if requested by either the participant or treating clinician and probed the potential influence of these medications on study findings in sensitivity analyses. Non-postpartum controls underwent rest and adenosine-stress myocardial perfusion PET with ^13^N-ammonia.^22^ Absolute rest and hyperemic MBF were calculated by fitting the ^13^N-ammonia or ^82^Rb time-activity curves to a validated two-compartment tracer kinetic model.^23^ MFR was calculated as the ratio of hyperemic to rest MBF. Coronary vascular resistance (CVR) was calculated as mean arterial pressure divided by MBF. Analysis was performed with blinding to preeclampsia status.

### Transthoracic Echocardiography

Echocardiography was performed on postpartum participants using GE E95 machines by experienced, licensed sonographers. Measures of left and right ventricular structure and function, left atrial size, and diastolic function were analyzed according to contemporary echocardiography guidelines.^25,26^ Additional details are provided in the **Supplemental Methods**.

### Angiogenic Biomarkers

The ratio of sFlt-1 to PlGF reflects angiogenic balance in pregnancy, is validated among pregnant individuals with suspected preeclampsia to “rule out” short-term progression to severe preeclampsia,^27^ and was recently approved by the U.S. Food and Drug Administration to guide obstetrical management. We assayed sFlt-1 and PlGF on venous blood serum drawn immediately prior to PET imaging using enzyme-linked immunosorbent assays (R&D Systems, Inc., Minneapolis, MN).

### Outcomes

The primary study outcome was MFR as measured by cardiac PET imaging. Secondary outcomes were MBF and CVR both at rest and with vasodilator stress.

### Statistical Analysis

Participant characteristics were compared across study groups using analysis of variance or the Kruskal-Wallis test for normally distributed and skewed continuous variables, respectively, and using the Fisher exact test for categorical variables. Primary models used unadjusted linear regression to calculate differences in PET indices (MFR, rest MBF, stress MBF, rest CVR, and stress CVR) among preeclamptic postpartum and normotensive postpartum participants vs. non-postpartum study participants (reference group). Given the older age of non-postpartum controls vs. postpartum participants, we additionally calculated differences in PET indices between groups with adjustment for age.

To probe robustness of our findings, we conducted sensitivity analyses that excluded women with pre-pregnancy chronic hypertension and gestational diabetes and further adjusted for BMI at the time of imaging. We additionally performed subgroup analyses that stratified women with preeclampsia (1) by delivery <34 weeks’ vs. ≥34 weeks’ gestation and (2) by whether women required antihypertensive medication on the morning of the study visit.

In exploratory analyses, we examined whether timing of imaging relative to delivery or clinical characteristics/routine laboratory biomarkers used in the diagnosis or monitoring of preeclampsia were associated with postpartum MFR, MBF, or CVR among individuals with preeclampsia using unadjusted linear regression. In addition, we compared echocardiographic parameters between women with preeclampsia and normotensive postpartum women (reference group) using the Student’s *t*-test or Wilcoxon rank-sum test, as appropriate. Pearson correlation coefficients tested the relationship of with MFR and MBF with echocardiographic indices among women with preeclampsia. In analyses of angiogenic biomarkers (sFlt-1, PlGF, and the sFlt-1/PlGF ratio) measured at the time of PET imaging, we also used Pearson correlation coefficients to test the relationship of log-transformed biomarkers with MFR, MBF, and CVR.

Sample size was calculated based on prior literature of flow-mediated dilation in women with preeclampsia^12^ and on available PET measurements in healthy female controls to provide 80% power to detect a 20% difference in MFR among those with preeclampsia vs. non-postpartum controls with a two-sided alpha of 0.05.

Two-sided *P*<0.05 indicated statistical significance; findings from secondary and exploratory analyses should be considered supportive and hypothesis-generating. Analyses were performed in R version 4.3.1.

## Results

The primary PET imaging cohort included 19 postpartum women with severe preeclampsia (mean [SD] age 32.9 [2.8] years, 15.3 [7.6] days postpartum at study assessment), 5 postpartum women following normotensive pregnancy (31.5 [3.6] years, 14.4 [8.4] days postpartum at assessment; **Figure S2**), and 13 non-postpartum pre-menopausal female controls (40.1 [8.5] years at assessment; **Table 1**). Overall, 27 of 37 participants (73.0%) were White, 5 (13.5%) were Black, 2 (5.4%) were Asian, and 3 (8.1%) were Hispanic. Of the 24 postpartum women studied, 15 (62.5%) were primiparous (i.e., had borne a total of one offspring) at the time of the study assessment, with a similar distribution of parity between those with preeclampsia and normotensive index pregnancies (**Table 1**). No participants had chronic diabetes, and 1 participant with preeclampsia also had gestational diabetes. Among those with preeclampsia in the index pregnancy, 4 (21.1%) had chronic pre-pregnancy hypertension with superimposed preeclampsia, 6 (31.6%) had used low-dose aspirin for preeclampsia prevention, and all but 1 (94.7%) delivered via Cesarean section. Participants with severe preeclampsia were enriched for delivery before 34 weeks’ gestation in the index pregnancy (14/19 [73.7%]), and gestational age at delivery was earlier in those with preeclampsia vs. normotensive pregnancy. Body mass index (BMI), SBP, DBP, and heart rate were each higher at the time of study assessments in women with preeclampsia vs. both normotensive postpartum individuals and non-postpartum controls (**Table 1**).

**Table 1.**
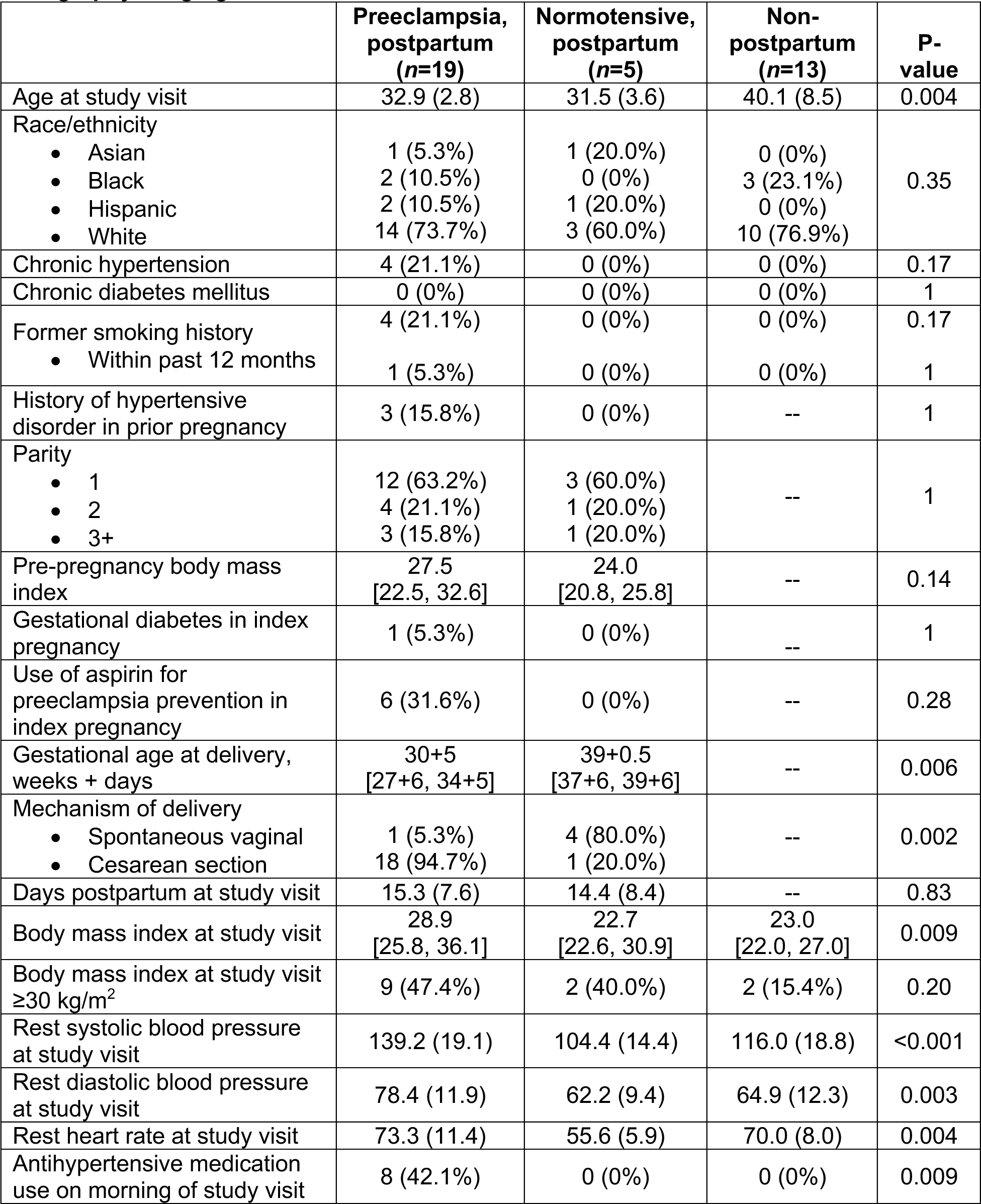
Characteristics of the primary analytic cohort with cardiac positron emission tomography imaging.

### Association of Preeclampsia with Myocardial Flow Reserve, Myocardial Blood Flow, and Coronary Vascular Resistance

The mean (SD) MFR was 2.82 (0.70) in women following delivery with preeclampsia, 3.14 (0.55) following normotensive pregnancy, and 3.49 (0.84) in non-postpartum controls. Compared with values in non-postpartum individuals, preeclampsia was associated with lower MFR (*β*=-0.67 [95% CI, −1.21 to −0.13]; *P*=0.016; **Figure 1**). The corresponding difference in individuals following normotensive pregnancy was −0.35 (95% CI, −1.14 to +0.44; *P*=0.37). After adjustment for age, preeclampsia and normotensive pregnancy were associated with differences in MFR of −0.76 (95% CI, −1.39 to −0.14; *P*=0.018) and −0.47 (95% CI, −1.34 to +0.41; *P*=0.29), respectively, vs. non-postpartum women (**Table 2**).

**Figure 1.**
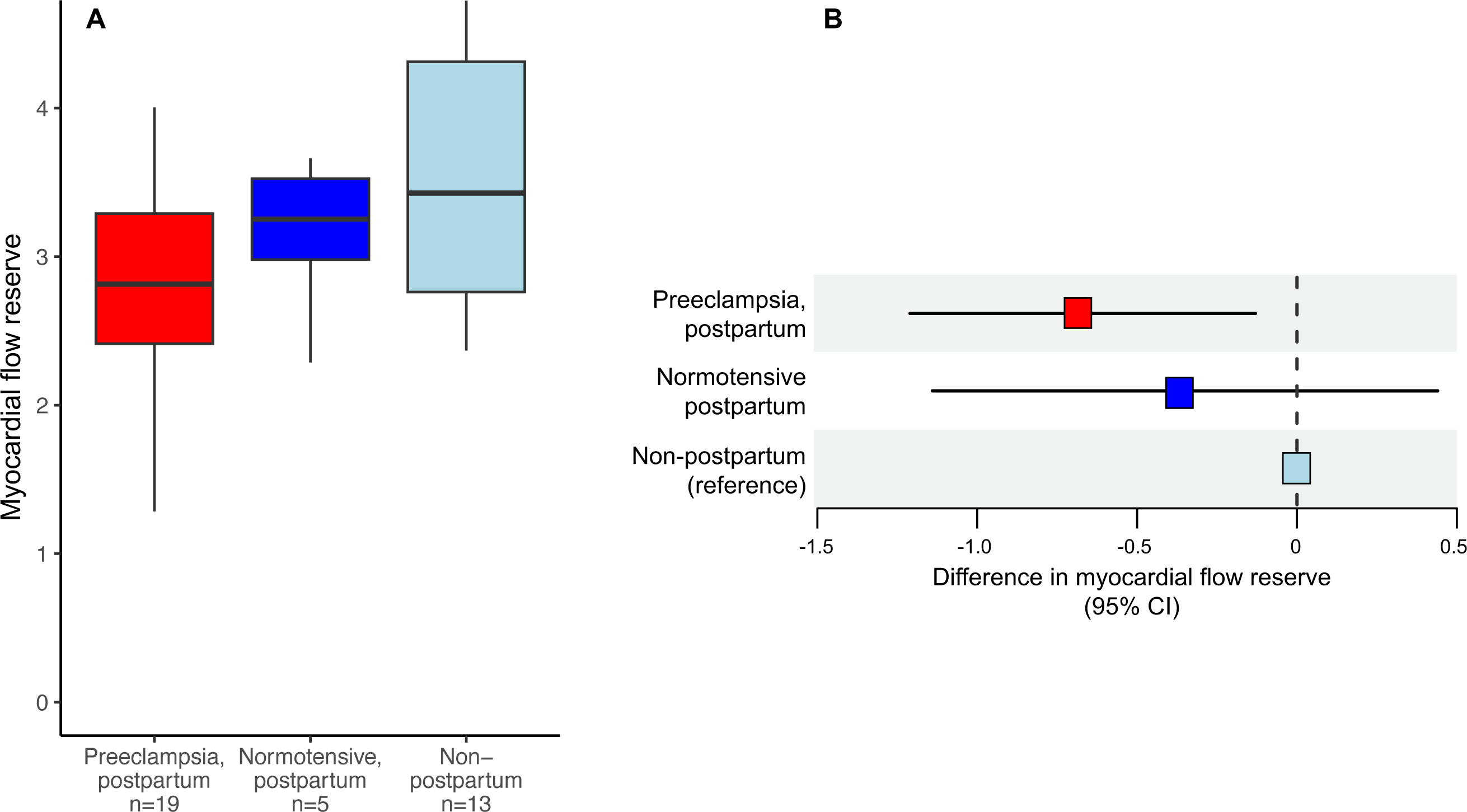
Severe preeclampsia is associated with reduced myocardial flow reserve in the early postpartum period. (A) Myocardial flow reserve by group. (B) Difference in myocardial flow reserve among postpartum women with severe preeclampsia and normotensive postpartum women vs. non-postpartum controls. Women with severe preeclampsia (*n*=19) underwent PET imaging at a mean (SD) 15.3 (7.6) days postpartum; normotensive postpartum women (*n*=5) underwent PET imaging at 14.4 (8.4) days postpartum. Non-postpartum women (*n*=13) constituted the reference group. Myocardial flow reserve was significantly reduced among women following delivery with severe preeclampsia.

**Table 2.** Myocardial flow reserve, myocardial blood flow, coronary vascular resistance, and differences vs. non-postpartum participants by group.

|  | Preeclampsia, postpartum<br>(n=19) |  |  |  |  | Normotensive, postpartum<br>(n=5) |  |  |  |  | Non-postpartum<br>(n=13) |
| --- | --- | --- | --- | --- | --- | --- | --- | --- | --- | --- | --- |
|  | Mean<br>(SD) | Unadjusted<br>difference vs.<br>non-postpartum |  | Age-adjusted<br>difference vs.<br>non-postpartum |  | Mean<br>(SD) | Unadjusted<br>difference vs.<br>non-postpartum |  | Age-adjusted<br>difference vs.<br>non-postpartum |  | Mean<br>(SD) |
|  |  | *Beta*<br>(95%<br>CI) | *P*-<br>value | *Beta*<br>(95%<br>CI) | *P*-<br>value |  | *Beta*<br>(95%<br>CI) | *P*-<br>value | *Beta*<br>(95%<br>CI) | *P*-<br>value |  |
| **Myocardial flow**<br>**reserve** | 2.82<br>(0.70) | -0.67<br>(-1.21<br>to -<br>0.13) | 0.016 | -0.76<br>(-1.39 to<br>-0.14) | 0.018 | 3.14<br>(0.55) | -0.35<br>(-1.14<br>to 0.44) | 0.37 | -0.47<br>(-1.34 to<br>0.41) | 0.29 | 3.49<br>(0.84) |
| **Rest**<br>**MBF,**<br>**mL/min/g** | 0.94<br>(0.29) | -0.02<br>(-0.19<br>to 0.16) | 0.85 | 0.02<br>(-0.17 to<br>0.22) | 0.81 | 0.73<br>(0.18) | -0.22<br>(-0.47<br>to 0.03) | 0.08 | -0.17<br>(-0.45 to<br>0.11) | 0.21 | 0.96<br>(0.15) |
| **Stress**<br>**MBF,**<br>**mL/min/g** | 2.55<br>(0.64) | -0.68<br>(-1.07<br>to -<br>0.29) | 0.001 | -0.61<br>(-1.06 to<br>-0.16) | 0.01 | 2.23<br>(0.21) | -1.01<br>(-1.58<br>to -<br>0.43) | 0.001 | -0.92<br>(-1.56 to<br>-0.28) | 0.006 | 3.24<br>(0.43) |
| **Rest**<br>**CVR,**<br>**mmHg/mL/min/g** | 110.9<br>(39.7) | 23.9<br>(1.3 to<br>46.5) | 0.04 | 22.6<br>(-3.7 to<br>48.9) | 0.09 | 94.0<br>(14.8) | 6.9<br>(-26.1<br>to 39.9) | 0.67 | 5.4<br>(-31.6 to<br>42.4) | 0.77 | 87.0<br>(16.0) |
| **Stress**<br>**CVR,**<br>**mmHg/mL/min/g** | 38.2<br>(10.8) | 12.4<br>(6.0 to<br>18.7) | <0.001 | 12.1<br>(4.7 to<br>19.5) | 0.002 | 32.2<br>(1.9) | 6.4<br>(-2.9 to<br>15.7) | 0.17 | 6.1<br>(-4.3 to<br>16.6) | 0.24 | 25.8<br>(6.2) |

MBF indicates myocardial blood flow. CVR indicates coronary vascular resistance. SD indicates standard deviation.

There were no significant differences in rest MBF in either the preeclampsia or normotensive postpartum groups vs. non-postpartum controls (**Table 2; Figure S3**). However, stress MBF was significantly reduced in both postpartum groups (preeclampsia: *β*=-0.68 [95% CI, −1.07 to −0.29] mL/min/g; *P*=0.001; normotensive: *β*=-1.01 [95% CI, −1.58 to −0.43] mL/min/g; *P*=0.001). Compared with non-postpartum individuals, women with preeclampsia had higher rest CVR (*β*=+23.9 [95% CI, 1.3 to 46.5] mmHg/mL/min/g; *P*=0.04) and stress CVR (*β*=+12.4 [95% CI, 6.0 to 18.7] mmHg/mL/min/g; *P*<0.001), with values observed in normotensive postpartum individuals falling between those of the preeclamptic and non-postpartum groups (**Table 2**).

Differences in PET indices among women following preeclampsia vs. non-postpartum controls were highly consistent in sensitivity analyses restricted to women without pre-pregnancy chronic hypertension or women without gestational diabetes (**Table S1**). In models further adjusted for BMI at the time of imaging, estimated differences in MFR were similar but estimated differences in stress MBF, rest CVR, and stress CVR for preeclamptic participants were mildly attenuated compared with primary analyses (**Table S2**). Findings were also broadly consistent in women with preeclampsia and delivery either before or after 34 weeks’ gestation, apart from a greater increase in rest CVR observed among those with delivery after 34 weeks (**Table S3**).

Among women with preeclampsia, MFR was positively associated with the number of days postpartum at the time of PET imaging (0.05 [95% CI, 0.02 to 0.09] per day; *P*=0.008; **Figure 3A**). Rest MBF decreased with the number of days postpartum (−0.02 [95% CI, −0.04 to − 0.004] per day; *P*=0.02; **Figure 3B**), whereas there was no apparent temporal relationship with stress MBF in the early postpartum period (**Table S4**).

**Figure 2.**
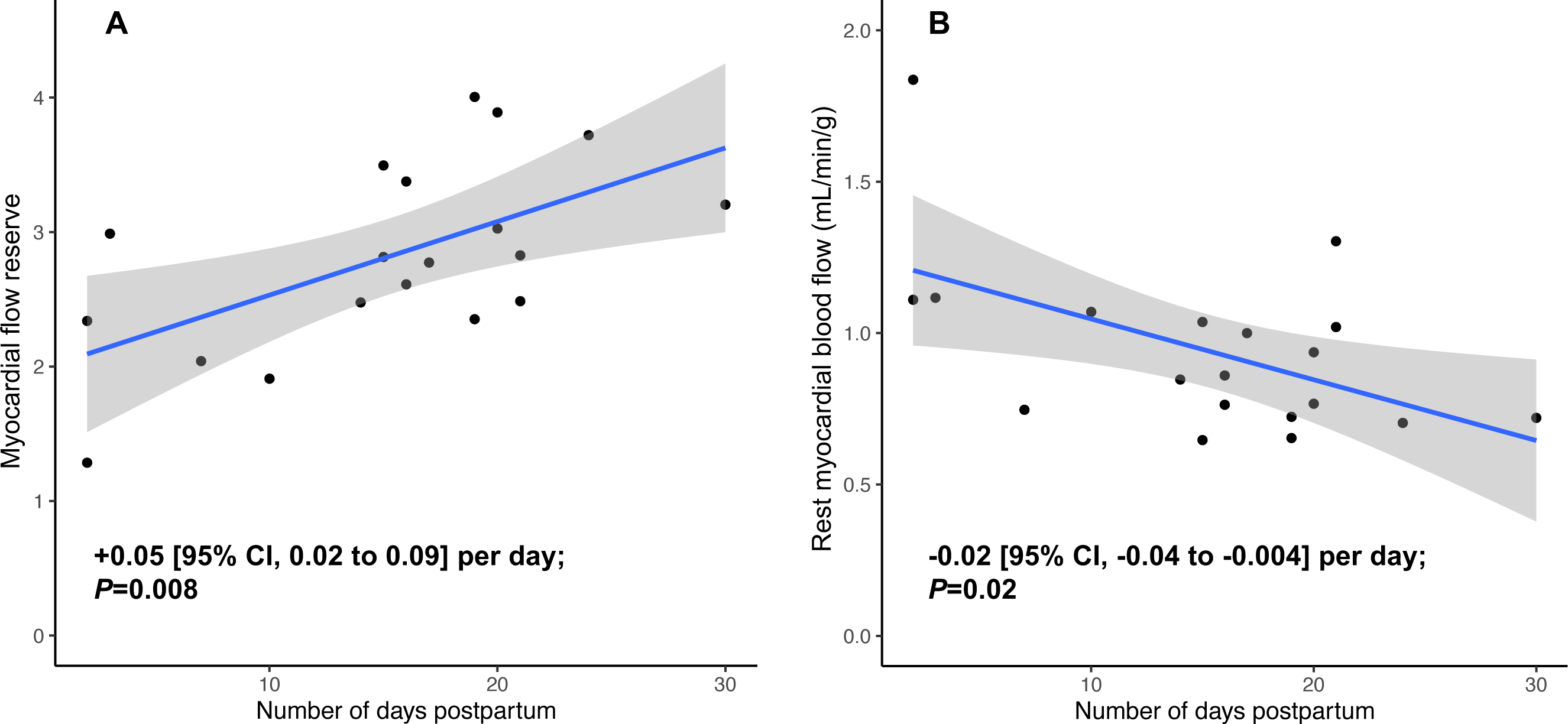
(A) Myocardial flow reserve and (B) rest myocardial blood flow vs. time following delivery among women with preeclampsia. Women with severe preeclampsia (*n*=19) underwent PET imaging at a mean (SD) 15.3 (7.6) days postpartum (overall range: 2-30 days). Myocardial flow reserve appeared to increase, and rest myocardial blood draw appeared to decrease, with time following delivery.

**Figure 3.**
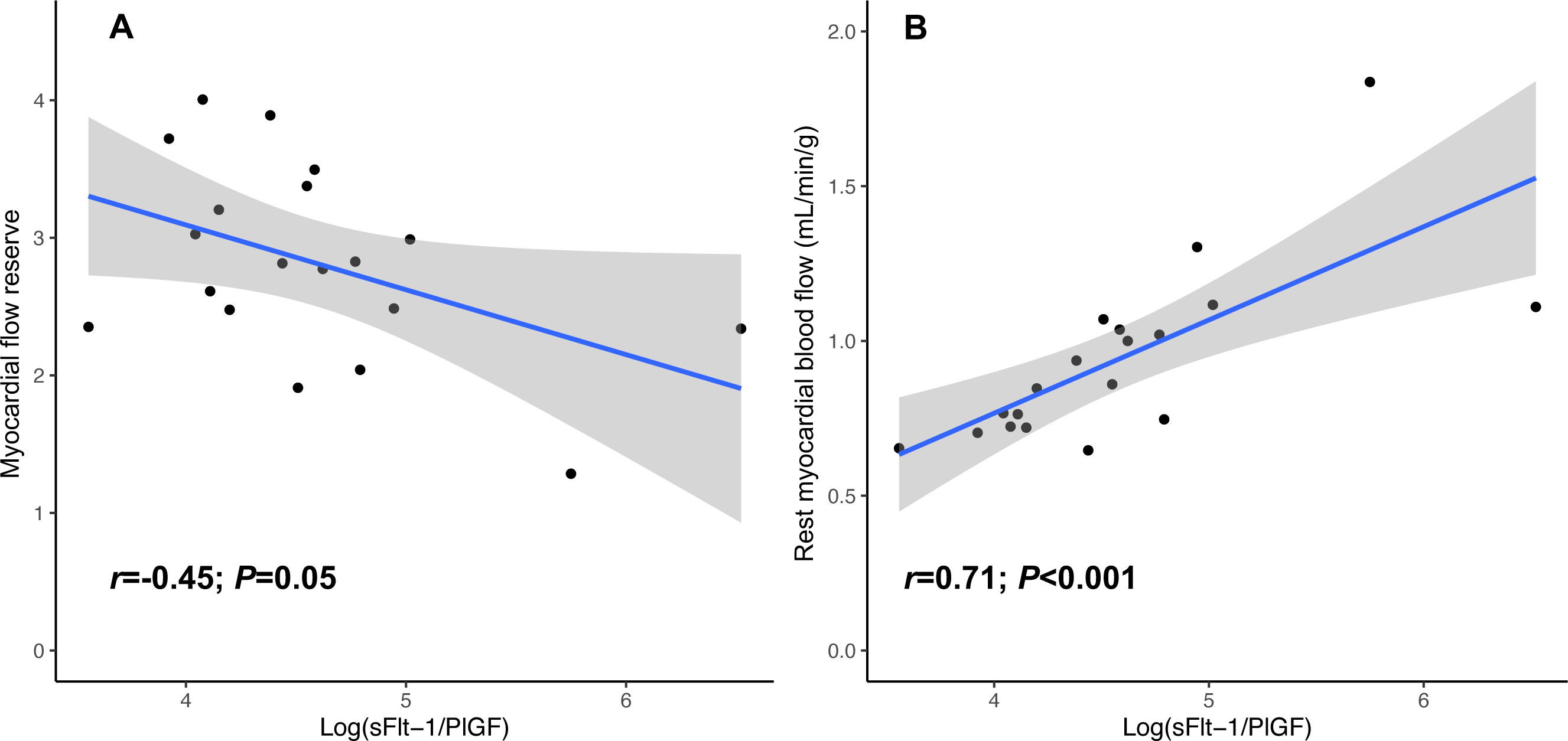
Correlation of (A) myocardial flow reserve and (B) rest myocardial blood flow with the sFlt-1/PlGF ratio among women with preeclampsia. Myocardial flow reserve was moderately inversely associated, and rest myocardial blood flow strongly positive correlated, with the sFlt-1/PlGF ratio among postpartum women with preeclampsia (*n*=19). sFlt-1 indicates soluble fms-like tyrosine kinase receptor-1. PlGF indicates placental growth factor.

Eight participants with preeclampsia (42.1%) required use of 1 or more antihypertensive medications on the morning of the study visit (7 used nifedipine, 4 used labetalol, and 1 used enalapril). Those who required antihypertensive medication completed study assessments closer to delivery than those who did not (median [IQR] 14.5 [8.3, 16.0] days postpartum vs. 19.0 [16.0, 20.5] days postpartum, respectively; *P*=0.10). Compared with non-postpartum participants, MFR was lower by 0.84 (95% CI 0.10-1.49; *P*=0.01) in those who required antihypertensive medication and lower by 0.55 (95% CI −0.12 to 1.22; *P*=0.10) in other participants with preeclampsia. Those who took antihypertensive medication on the morning of the study visit had attenuated differences in rest CVR compared with those who did not but had similar differences in rest MBF, stress MBF, and stress CVR (**Table S5**).

### Clinical Correlates of Postpartum PET Indices in Women with Severe Preeclampsia

Maximum recorded antepartum SBP was nominally associated with reduced stress MBF at postpartum PET imaging (*β*=-0.02 [95% CI, −0.03 to −0.001] mL/min/g per mmHg SBP; *P*=0.04). Maximum recorded DBP, maximum recorded proteinuria, and other laboratory biomarkers used to diagnose preeclampsia and its subtypes were each not significantly associated with postpartum MFR or other PET indices (**Table S6**). Report of headache as part of preeclampsia symptomatology vs. no reported headache was associated with reduced stress MBF (*β*=-0.66 [95% CI −1.21 to −0.12] mL/min/g; *P*=0.02) and increased stress CVR (*β*=+11.8 [95% CI 2.8 to 20.9] mmHg/mL/g/min; *P*=0.01). BMI ≥30 kg/m^2^ vs. <30 kg/m^2^ at the time of PET imaging was associated with reduced stress MBF (*β*=-0.72 [95% CI −1.24 to −0.20] mL/min/g; *P*=0.01), increased rest CVR (*β*=+48.1 [95% CI 17.1 to 79.2] mmHg/mL/g/min; *P*=0.02), and increased stress CVR (*β*=+14.9 [95% CI 7.3 to 22.5] mmHg/mL/g/min; *P*<0.001) (**Table S6**). Hematocrit nadir during the delivery admission (mean [SD] 28.3 [4.0]%) was not associated with any postpartum PET indices.

### Postpartum Echocardiographic Differences Between Groups and PET Correlates in Women with Severe Preeclampsia

Transthoracic echocardiography was performed in 17 of the 19 enrolled postpartum women with preeclampsia (mean [SD] 15.5 [7.9] days postpartum at study assessment) and in 7 women following normotensive pregnancy (13.7 [7.8] days postpartum at assessment; **Figure S1**; **Table S7**). Compared with normotensive postpartum women, those with preeclampsia had greater LV wall thickness (mean [SD] septal wall thickness: 8.9 [2.0] vs. 6.7 [0.4] mm; *P*<0.001; posterior wall thickness: 9.1 [1.5] vs. 6.8 [1.0] mm; *P*<0.001) and greater relative wall thickness (0.38 [0.07] vs. 0.30 [0.07]; *P*=0.03). LV ejection fraction did not differ between groups (**Table 3**). In addition, women with preeclampsia had reduced lateral e’ velocity (13.7 [2.4] vs. 16.8 [2.1] cm/s; *P*=0.008) and higher E/e’ (median [IQR] 7.8 [6.6, 8.0] vs. 5.7 [5.0, 7.2]; *P*=0.04). Mean (SD) left atrial volume was 55.7 (12.6) mL in participants with preeclampsia vs. 45.5 (10.5) mL in those with normotensive pregnancy (*P*=0.06). Regional and global LV strain indices were numerically higher in women with preeclampsia vs. normotensive pregnancy, although differences did not reach statistical significance (**Table 3**).

**Table 3.** Echocardiographic findings in postpartum participants.

|  | **Preeclampsia,**<br>**postpartum**<br>**(n=17)** | **Normotensive,**<br>**postpartum**<br>**(n=7)** | **P-value** |
| --- | --- | --- | --- |
| Septal wall, mm | 8.9 (2.0) | 6.7 (0.4) | <0.001 |
| Posterior wall, mm | 9.1 (1.5) | 6.8 (1.0) | <0.001 |
| Relative wall thickness | 0.38 (0.07) | 0.30 (0.07) | 0.03 |
| LV end-diastolic dimension, mm | 47.7 (4.0) | 46.0 (6.2) | 0.52 |
| LV end-systolic dimension, mm | 33.9 (4.3) | 31.8 (4.2) | 0.29 |
| LV ejection fraction, % | 57.2 (4.1) | 57.8 (3.8) | 0.75 |
| LA volume, mL | 55.7 (12.6) | 45.5 (10.5) | 0.06 |
| Mitral E, cm/s | 84.5 (13.1) | 82.6 (13.7) | 0.76 |
| Mitral A, cm/s | 58.1 (10.9) | 51.2 (17.9) | 0.37 |
| Mitral E/A ratio | 1.4 [1.4, 1.7] | 2.0 [1.3, 2.0] | 0.38 |
| Lateral e', cm/s | 13.7 (2.6) | 16.8 (2.1) | 0.009 |
| Septal e', cm/s | 11.0 (2.7) | 12.2 (2.9) | 0.35 |
| Average E/e' ratio | 7.8 [6.6, 8.0] | 5.7 [5.0, 7.2] | 0.04 |
| Tricuspid regurgitant velocity, cm/s | 1.9 (0.4) | 1.7 (0.2) | 0.18 |
| Tricuspid annular S', cm/s | 12.5 (2.3) | 12.7 (2.2) | 0.86 |
| Tricuspid annular plane systolic excursion, cm | 23.9 (4.7) | 22.5 (3.7) | 0.45 |
| RV fractional area change, % | 42.3 (5.3) | 40.3 (5.8) | 0.45 |
| Basal LV longitudinal strain, % | -14.2 (3.1) | -16.2 (2.3) | 0.10 |
| Mid-LV longitudinal strain, % | -14.3 (3.8) | -15.5 (1.8) | 0.32 |
| Apical LV longitudinal strain, % | -14.2 (4.8) | -17.1 (4.8) | 0.21 |
| Global LV longitudinal strain, % | -14.3 (3.5) | -16.3 (2.4) | 0.12 |

Values are displayed as mean (SD) or median [IQR] for normally distributed and skewed variables, respectively. LV indicates left ventricular. RV indicates right ventricular.

In exploratory analyses, we did not observe any significant correlations between MFR and echocardiographic parameters among women with severe preeclampsia (**Table S8**).

### Postpartum Angiogenic Factor Levels and PET Indices in Women with Severe Preeclampsia

Among women with preeclampsia, the median [IQR] sFlt-1 level was 435.5 [388.6, 582.4] pg/mL, the PlGF level was 6.1 [5.2, 7.3] pg/mL, and the sFlt-1/PlGF ratio was 90.8 [62.2, 119.2] at the time of PET imaging. The sFlt-1/PlGF ratio declined exponentially with time postpartum (**Figure S4**). The sFlt-1/PlGF ratio was moderately inversely correlated with MFR (*r*=-0.45; *P*=0.05; **Figure 4A**; **Table S9**), driven by a stronger positive correlation with rest MBF (*r*=0.71; *P*<0.001; **Figure 4B**) than with stress MBF (*r*=0.22; *P*=0.36). These correlations were stronger after excluding participants who required antihypertensive medication on the morning of imaging assessment (MFR: *r*=-0.80, *P*=0.003; rest MBF: *r*=0.91, *P*<0.001; **Figure S5**). In a *post hoc* model mutually adjusted for both rate-pressure product and the sFlt-1/PlGF ratio, only the sFlt-1/PGF ratio was independently associated with rest MBF (*β*=0.21 [95% CI, 0.05 to 0.37] mL/min/g per log-unit; *P*=0.01).

## Discussion

Coronary microvascular dysfunction has been hypothesized to underlie both acute and longer-term cardiovascular morbidity in women with preeclampsia. To our knowledge, this analysis represents the first interrogation of coronary blood flow and microvascular reactivity in the peripartum period following preeclampsia. In line with our hypothesis, we observed a reduction in the maximum adenosine-stimulated flow response and a related increase in CVR following severe preeclampsia vs. the non-postpartum state. Furthermore, MFR appeared to recover with time postpartum following preeclampsia due to normalization of rest flows; this normalization of rest flow was tightly correlated with the sFlt-1/PlGF ratio, independent of hemodynamics. Of note, normotensive postpartum participants also had significantly decreased stress MBF and intermediate findings with respect to MFR, suggesting that a portion of observed differences may reflect normal postpartum physiology that is compounded or exaggerated in the context of preeclampsia. Taken together, these findings support the notion that preeclampsia exerts adverse coronary microvascular effects and may have implications for understanding risk of short-term (e.g., PPCM) and long-term cardiovascular complications in affected women.

First, our findings reinforce the relevance of the microcirculation in preeclampsia. SFlt-1 is believed to exert its anti-angiogenic effects by binding to and blocking the pro-angiogenic properties of PlGF and VEGF.^1^ In preclinical models, sFlt-1 blocks angiogenesis-associated migration of endothelial cells^14^ and provokes constriction of rat renal arterioles.^11^ The anti-angiogenic effects of sFlt-1 may represent a “second hit” triggering PPCM in susceptible individuals and may therefore explain this condition’s peak incidence in the very early postpartum period, immediately after circulating anti-angiogenic factor levels have peaked.^14,28^ A recent investigation further reported that mice with sFlt-1 overexpression in pregnancy demonstrated enhanced mesenteric vasoconstriction vs. wild-type mice in response to an angiotensin II challenge at two months postpartum, implying potential enduring effects on sensitivity to pro-hypertensive stimuli beyond the peripartum period.^29^ Furthermore, human studies have suggested microvascular rarefaction in the retina^30^ and fingers^31^ of midlife women with a history of preeclampsia, independent of blood pressure. Collectively, these findings imply potential “microvascular susceptibility” in women with preeclampsia. Although our findings suggested short-term improvement in MFR related to normalization of rest MBF with time postpartum, ongoing coronary microvascular susceptibility following preeclampsia may contribute to the excess risk of HFpEF^18^ reported in affected women.^7,9^ Future longitudinal studies will enable further interrogation of this hypothesis.

Second, placental biomarkers may provide insights regarding how preeclampsia affects myocardial blood flow. Among women with severe preeclampsia in our study, the sFlt-1/PlGF ratio correlated strongly with rest MBF. Furthermore, the correlation of sFlt-1/PlGF with rest MBF among preeclamptic women was not explained by the rate-pressure product, suggesting that preeclampsia exerts hemodynamic-independent effects on cardiomyocyte oxygen utilization. Through feedback between cardiomyocytes and the coronary microcirculation, coronary arterioles may dilate to increase MBF in the resting state to compensate for impaired tissue oxygen extraction and/or utilization. Prior work demonstrates impaired oxygen utilization in the context of severe preeclampsia.^32,33^ The association of the sFlt-1/PlGF ratio with increased rest MBF may therefore reflect an effort by cardiomyocytes to overcome microvascular and/or cardiomyocyte dysfunction driven by circulating placental proteins.

Third, our findings highlight key gaps in understanding of cardiac effects in normal pregnancy and normal postpartum cardiovascular adaptation. Reduction in stress MBF in both hypertensive and normotensive postpartum groups vs. non-postpartum women was an unexpected finding of this study which requires validation. Although this study was not powered to draw definitive conclusions about the normotensive postpartum group, these participants demonstrated reduction in stress MBF and an “intermediate phenotype” between those with preeclampsia and non-postpartum controls with respect to MFR and CVR. These findings raise the possibility that even uncomplicated pregnancy affects coronary microcirculatory function, a hypothesis that warrants further dedicated study.

Fourth, studies involving advanced cardiac imaging can be adapted to the peripartum period. Preterm preeclampsia was overrepresented in our cohort, partly reflecting greater willingness to participate in hospital-based research among women whose preterm infants were receiving care in the neonatal intensive care unit, thereby removing a key logistical barrier (need for childcare) to study participation. As sFlt-1 and the sFlt-1/PlGF ratio are generally higher at term than earlier in gestation even among those with preeclampsia,^34^ it is theoretically possible that the resulting gestational age imbalance served to minimize apparent differences between preeclamptic and normotensive participants. This experience highlights key challenges to conducting research involving new mothers and suggests potential value of protocol modifications (e.g., use of vasodilators with a shorter half-life such as adenosine) to minimize interruption of breastfeeding and of offering childcare or other supports to facilitate research participation during a critical period when the primary focus is appropriately on caring for the newborn.

### Limitations

This study should be considered in the context of limitations. First, given the novel application of PET imaging to young adults in the early postpartum period, we enrolled a sample size sufficient to test differences in the primary endpoint (MFR) among those with severe preeclampsia but not those with normotensive pregnancy, whose findings we expected *a priori* to be normal. Despite the modest sample size included, significant echocardiographic differences between preeclamptic and normotensive postpartum participants were detected and were consistent with the previous literature.^13^ Second, due to practical considerations of coordinating cardiac PET imaging in newly postpartum individuals, study visits occurred at an average of two weeks postpartum; differences among groups may have already partially attenuated by the time of study assessments. Although strictly standardized timing of PET imaging is not feasible in the newly postpartum population, timing of study visits was consistent between preeclamptic and normotensive postpartum groups, and assessments across the first four postpartum weeks enabled us to examine trends over time. Third, to ensure safety of preeclamptic participants, we permitted participants to use antihypertensive medications prior to study assessments if necessary, which may have influenced results. However, prior literature suggests these medications would be expected to have favorable effects on MFR,^35,36^ whereas those requiring antihypertensive medication in our study had evidence of greater reduction in MFR, possibly reflecting unresolved preeclampsia pathophysiology. Fourth, obstetrical history was not available in the non-postpartum control cohort, although no participants in this control cohort had chronic hypertension. Fifth, the postpartum groups were studied using a more recent generation PET camera than non-postpartum controls. However, all PET cameras used in the study were calibrated quarterly to ensure accurate activity concentration in PET images; therefore, measurements of arterial and tissue activity concentrations are expected to be consistent and reproducible across PET cameras. Finally, this study employed a cross-sectional design, and we cannot exclude the possibility that pre-conception MFR and MBF differ in those who go on to develop preeclampsia; future longitudinal studies will be required to clarify lifecourse trajectories of microvascular function.

## Perspectives

Overall, we found evidence of reduced coronary microvascular function in the postpartum period among individuals delivering with severe preeclampsia. These observations in humans corroborate previous findings in preclinical models and lend support to the notion that systemic microvascular dysfunction in preeclampsia also involves the coronary microcirculation. These findings may be relevant to preeclampsia-associated cardiovascular risk, including increased risk of heart failure. Future research is needed to clarify lifecourse trajectories of coronary and extracoronary microvascular function in women with preeclampsia and establish effective interventions to mitigate risk of preeclampsia-associated cardiovascular disease.

## Novelty and Relevance

### What is New?

- Among women with severe preeclampsia undergoing cardiac positron emission tomography perfusion imaging in the early postpartum period, myocardial flow reserve and stress myocardial blood flow were lower, and stress coronary vascular resistance was higher, compared with values in non-postpartum controls.
- Myocardial flow reserve appeared to recover with time following severe preeclampsia due to normalization of rest flows; this normalization of rest flow was tightly correlated with the sFlt-1/PlGF ratio and was independent of hemodynamics.

### What is Relevant?

- Our findings support the notion that systemic microvascular dysfunction in preeclampsia also involves the coronary microcirculation.

### Clinical/Pathophysiological Implications?

- Interventions that promote microvascular health may reduce the risk of preeclampsia-associated cardiovascular disease.
- Further research is needed to understand cardiac effects of normal pregnancy and their underlying mechanisms.

## Supplemental Material

Supplemental Methods

Figures S1-S5

Tables S1-S9

## Data Availability

All data produced in the present study are available upon reasonable request to the authors.

## Abbreviations and Acronyms

BMI: body mass index
CAD: coronary artery disease
CVR: coronary vascular resistance
HFpEF: heart failure with preserved ejection fraction
MBF: myocardial blood flow
MFR: myocardial flow reserve
PET: positron emission tomography
PlGF: placental growth factor
PPCM: peripartum cardiomyopathy
sFlt-1: soluble fms-like tyrosine kinase receptor 1

