## Supplemental Material for "Coronary Microvascular Function Following Severe Preeclampsia"

Supplemental Methods. 2

Figure S1. Participants included in analyses of cardiac positron emission tomography and echocardiographic indices. 3

Figure S2. Timing of study assessments among postpartum participants. 4

Figure S3. Differences in myocardial blood flow and coronary vascular resistance by group. Rest and stress coronary vascular resistance were increased in women following severe preeclampsia. Stress myocardial blood flow was reduced in both preeclamptic and normotensive postpartum women. 5

Figure S4. The ratio of sFlt-1/PlGF vs. days since delivery among women with preeclampsia. 6

Figure S5. Correlation of (a) myocardial flow reserve and (b) rest myocardial blood flow with the sFlt-1/PlGF ratio among women with preeclampsia who did not require antihypertensive medication prior to study imaging. 7

Table S1. Sensitivity analyses: Myocardial flow reserve, myocardial blood flow, coronary vascular resistance, and differences vs. non-postpartum individuals (a) excluding those with chronic hypertension and (b) excluding those with gestational diabetes. 8

Table S2. Sensitivity analysis: Myocardial flow reserve, myocardial blood flow, coronary vascular resistance, and differences vs. non-postpartum participants by group, with adjustment for age and body mass index at the time of imaging. 9

Table S3. Subgroup analyses: Myocardial flow reserve, myocardial blood flow, coronary vascular resistance, and differences vs. non-postpartum individuals in preeclampsia stratified by delivery before vs. at or after 34 weeks' gestation. 10

Table S4. Association of time following delivery with myocardial flow reserve, myocardial blood flow, and coronary vascular resistance among postpartum individuals with preeclampsia. 11

Table S5. Subgroup analyses: Myocardial flow reserve, myocardial blood flow, coronary vascular resistance, and differences vs. non-postpartum individuals in preeclampsia with or without use of antihypertensive medication prior to imaging. 12

Table S6. Association of clinical factors with PET indices among women with preeclampsia. 13

Table S7. Characteristics of the study cohort with echocardiographic data. 16

Table S8. Correlation of myocardial flow reserve, myocardial blood flow, and coronary vascular resistance with echocardiographic parameter among postpartum individuals with preeclampsia. 17

Table S9. Correlation of angiogenic biomarkers with PET indices. 18

**Supplemental Methods.**

*Medical History and Details of the Index Pregnancy*

Obstetrical and other medical history was collected by questionnaire administered by study staff and by review of the medical record. Among women with preeclampsia, clinical variables used to ascertain diagnosis, severity, and subtype of preeclampsia were extracted systematically from the medical record, including maximum antepartum systolic and diastolic blood pressures (SBP and DBP) and accompanying heart rate, maximum recorded proteinuria from a 24-hour urine collection or as estimated by a protein-to-creatinine ratio (standardized as equivalents of grams per 24 h for analysis), headache, other neurologic symptoms, pulmonary edema confirmed by chest radiography, and indices of kidney and liver function.

*Transthoracic Echocardiography*

Echocardiographic indices of interest were left ventricular (LV) wall thickness and relative wall thickness, LV end-systolic and diastolic dimensions, left atrial volume, mitral inflow E and A velocities and the E/A ratio, septal and lateral e’ velocities and the average E/e’ ratio, tricuspid regurgitant jet velocity, tricuspid annular S’ velocity, tricuspid annular plane systolic excursion, and regional and global longitudinal strain. Relative wall thickness was calculated as 2*posterior wall thickness/LV end-diastolic dimension. LV ejection fraction and left atrial volume were each calculated using the biplane method of discs. Speckle tracking echocardiography software (Image-Arena version 4.6.6.3, TomTec, Munich, Germany) was used to measure LV longitudinal strain from apical two-, three-, and four-chamber views, segmented into basal, mid-ventricular, and apical segments. Interpretation of each echocardiogram was performed independently by two investigators blinded to case-control status, clinical characteristics, and prior imaging, and averaged values were used in analysis.

**Figure S1. Participants included in analyses of cardiac positron emission tomography and echocardiographic indices.**

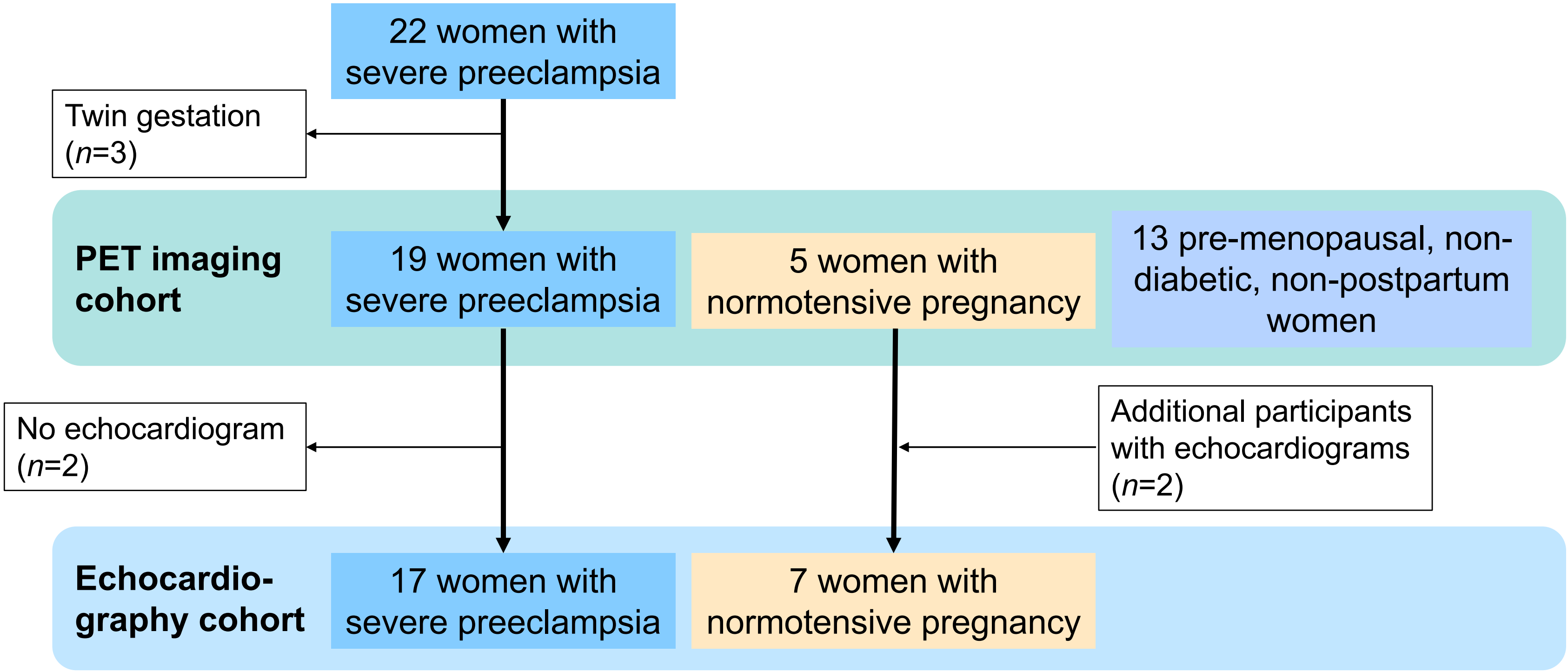

**Figure S2. Timing of study assessments among postpartum participants.**

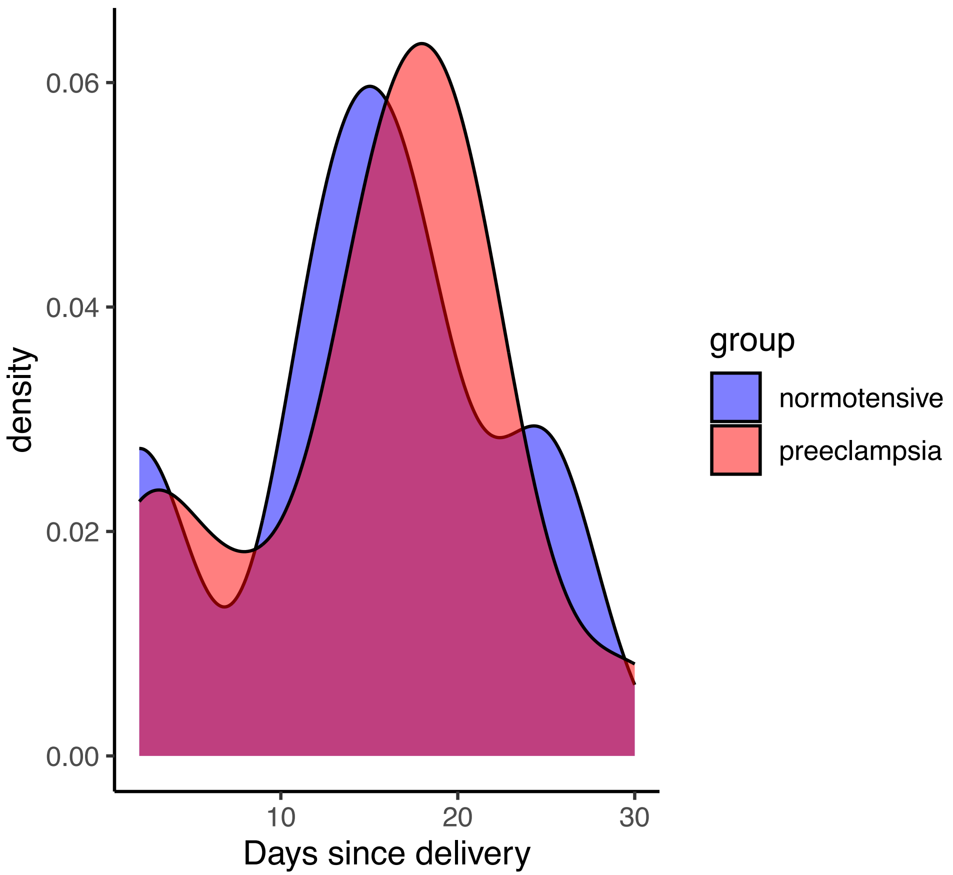

**Figure S3. Differences in myocardial blood flow and coronary vascular resistance by group.** Rest and stress coronary vascular resistance were increased in women following severe preeclampsia. Stress myocardial blood flow was reduced in both preeclamptic and normotensive postpartum women.

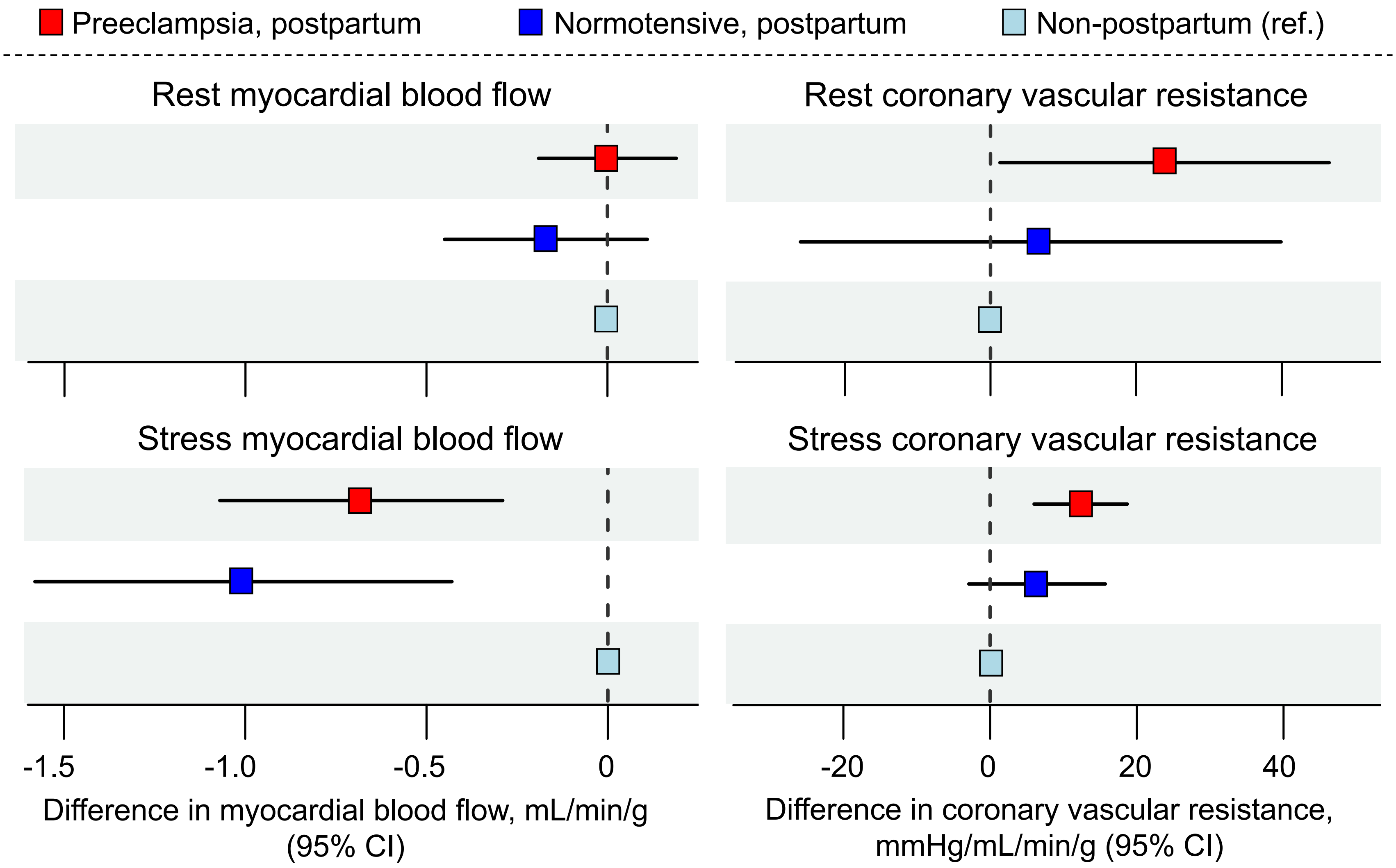

**Figure S4. The ratio of sFlt-1/PlGF vs. days since delivery among women with preeclampsia (*n*=19).**

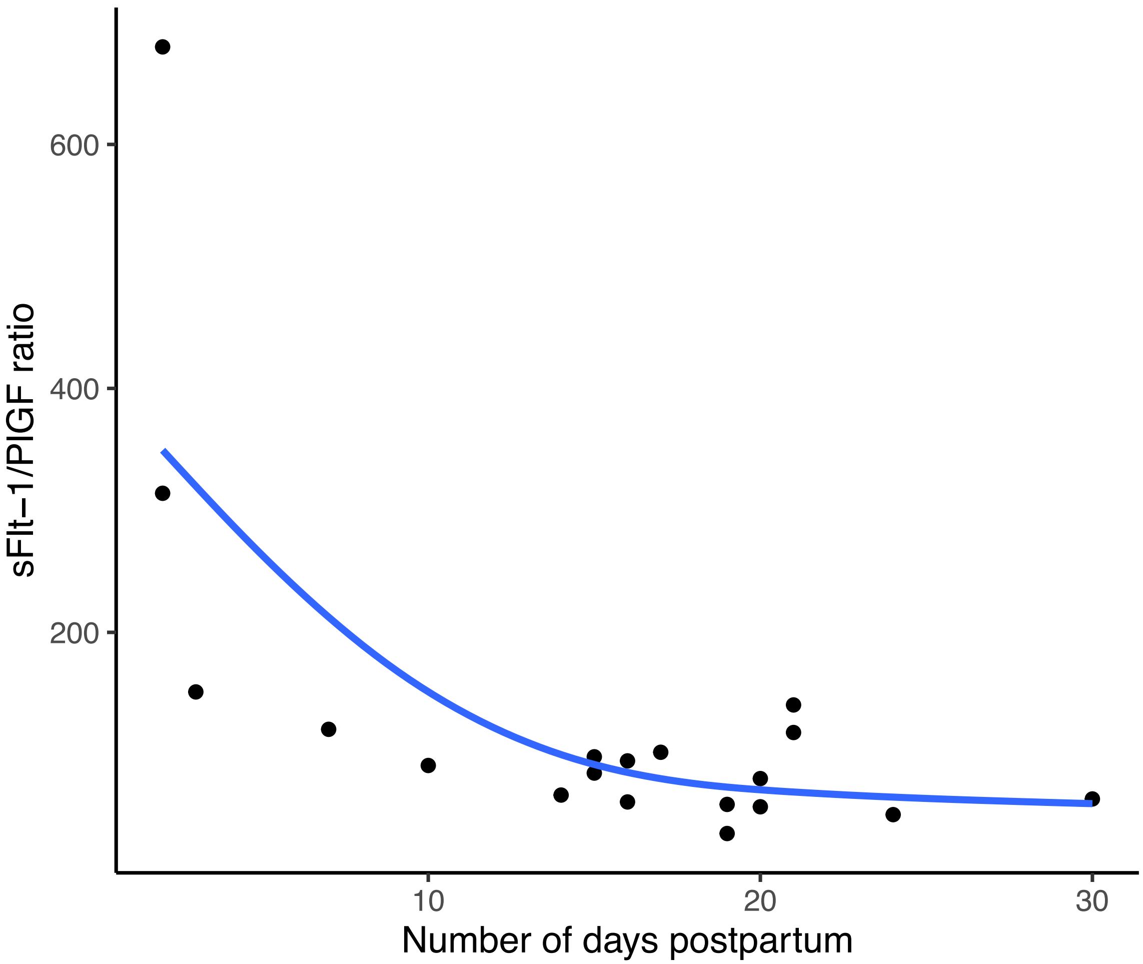


Data were fit using a generalized additive model in the *ggplot2* package (R version 4.3.1). sFlt-1 indicates soluble fms-like tyrosine kinase receptor-1. PlGF indicates placental growth factor.

**Figure S5. Correlation of (a) myocardial flow reserve and (b) rest myocardial blood flow with the sFlt-1/PlGF ratio among women with preeclampsia who did not require antihypertensive medication prior to study imaging (*n*=11).** sFlt-1 indicates soluble fms-like tyrosine kinase receptor-1. PlGF indicates placental growth factor.

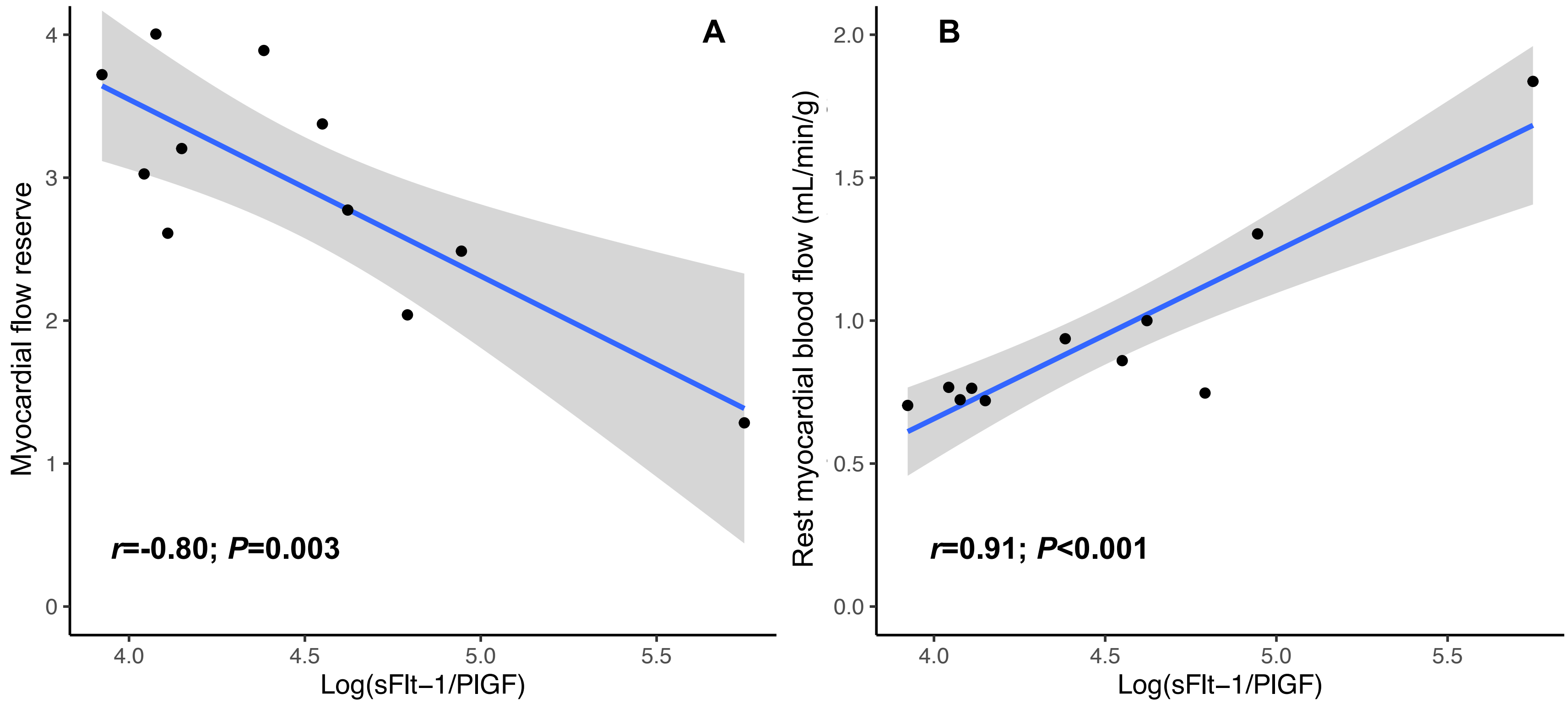

**Table S1. Sensitivity analyses: Myocardial flow reserve, myocardial blood flow, coronary vascular resistance, and differences vs. non-postpartum individuals (a) excluding those with chronic hypertension and (b) excluding those with gestational diabetes.**

| | **(a) Preeclampsia, excluding pre-pregnancy chronic hypertension (*n*=15 included)** | | | **(b) Preeclampsia, excluding gestational diabetes (*n*=18 included)** | | | **Non-post-partum (*n*=13)** |
|---|---|---|---|---|---|---|---|
| | Mean (SD) | Difference vs. non-postpartum | | Mean (SD) | Difference vs. non-postpartum | | Mean (SD) |
| | | *Beta (95% CI)* | *P-value* | | *Beta (95% CI)* | *P-value* | |
| **Myocardial flow reserve** | 2.81 (0.76) | -0.68 (-1.28 to -0.09) | 0.03 | 2.84 (0.72) | -0.65 (-1.20 to -0.10) | 0.02 | 3.49 (0.84) |
| **Rest MBF**, mL/min/g | 0.95 (0.31) | 0.00 (-0.19 to 0.19) | 0.97 | 0.95 (0.29) | -0.01 (-0.19 to 0.16) | 0.90 | 0.96 (0.15) |
| **Stress MBF**, mL/min/g | 2.53 (0.55) | -0.70 (-1.07 to -0.34) | <0.001 | 2.58 (0.65) | -0.66 (-1.06 to -0.26) | 0.002 | 3.24 (0.43) |
| **Rest CVR**, mmHg/mL/min/g | 109.2 (41.7) | 22.2 (-1.5 to 46.0) | 0.07 | 111.4 (40.8) | 24.4 (1.2 to 47.6) | 0.04 | 87.0 (16.0) |
| **Stress CVR**, mmHg/mL/min/g | 37.7 (9.2) | 11.9 (6.1 to 17.7) | <0.001 | 37.7 (10.9) | 11.9 (5.4 to 18.3) | <0.001 | 25.8 (6.2) |

MBF indicates myocardial blood flow. CVR indicates coronary vascular resistance. SD indicates standard deviation. Differences between preeclampsia and non-postpartum groups were calculated using unadjusted linear regression.

**Table S2. Sensitivity analysis: Myocardial flow reserve, myocardial blood flow, coronary vascular resistance, and differences vs. non-postpartum participants by group, with adjustment for age and body mass index at the time of imaging.**

| | **Preeclampsia, postpartum (*n*=19)** | | | **Normotensive, postpartum (*n*=5)** | | | **Non-post-partum (*n*=13)** |
|---|---|---|---|---|---|---|---|
| | Mean (SD) | Adjusted difference vs. non-postpartum | | Mean (SD) | Adjusted difference vs. non-postpartum | | Mean (SD) |
| | | *Beta (95% CI)* | *P-value* | | *Beta (95% CI)* | *P-value* | |
| **Myocardial flow reserve** | 2.82 (0.70) | -0.68 (-1.39 to 0.02) | 0.056 | 3.14 (0.55) | -0.45 (-1.34 to 0.44) | 0.31 | 3.49 (0.84) |
| **Rest MBF**, mL/min/g | 0.94 (0.29) | 0.08 (-0.14 to 0.30) | 0.48 | 0.73 (0.18) | -0.16 (-0.44 to 0.11) | 0.24 | 0.96 (0.15) |
| **Stress MBF**, mL/min/g | 2.55 (0.64) | -0.39 (-0.87 to 0.09) | 0.11 | 2.23 (0.21) | -0.89 (-1.50 to -0.27) | 0.006 | 3.24 (0.43) |
| **Rest CVR**, mmHg/mL/min/g | 110.9 (39.7) | 6.2 (-20.4 to 32.8) | 0.64 | 94.0 (14.8) | 2.8 (-30.8 to 36.5) | 0.87 | 87.0 (16.0) |
| **Stress CVR**, mmHg/mL/min/g | 38.2 (10.8) | 7.1 (-0.2 to 14.5) | 0.058 | 32.2 (1.9) | 5.3 (-4.0 to 14.6) | 0.25 | 25.8 (6.2) |

MBF indicates myocardial blood flow. CVR indicates coronary vascular resistance. SD indicates standard deviation. Differences between each postpartum group and the non-postpartum group were calculated using multivariable-adjusted linear regression.

**Table S3. Subgroup analyses: Myocardial flow reserve, myocardial blood flow, coronary vascular resistance, and differences vs. non-postpartum individuals in preeclampsia stratified by delivery before vs. at or after 34 weeks' gestation.**

| | **Preeclampsia with delivery before 34 weeks (*n*=14)** | | | **Preeclampsia with delivery at or after 34 weeks (*n*=5)** | | | **Non-post-partum (*n*=13)** |
|---|---|---|---|---|---|---|---|
| | Mean (SD) | Difference vs. non-postpartum | | Mean (SD) | Difference vs. non-postpartum | | Mean (SD) |
| | | *Beta (95% CI)* | *P-value* | | *Beta (95% CI)* | *P-value* | |
| **Myocardial flow reserve** | 2.79 (0.57) | -0.71 (-1.29 to -0.12) | 0.02 | 2.92 (1.07) | -0.57 (-1.37 to 0.23) | 0.15 | 3.49 (0.84) |
| **Rest MBF**, mL/min/g | 0.91 (0.19) | -0.05 (-0.23 to 0.14) | 0.60 | 1.03 (0.48) | 0.07 (-0.18 to 0.32) | 0.56 | 0.96 (0.15) |
| **Stress MBF**, mL/min/g | 2.52 (0.70) | -0.71 (-1.14 to -0.29) | 0.002 | 2.64 (0.51) | -0.59 (-1.18 to -0.01) | 0.04 | 3.24 (0.43) |
| **Rest CVR**, mmHg/mL/min/g | 104.6 (30.2) | 17.6 (-6.1 to 41.3) | 0.14 | 128.5 (60.1) | 41.5 (9.1 to 73.9) | 0.01 | 87.0 (16.0) |
| **Stress CVR**, mmHg/mL/min/g | 38.2 (11.0) | 12.4 (5.4 to 19.3) | <0.001 | 38.2 (11.5) | 12.4 (2.9 to 21.9) | 0.01 | 25.8 (6.2) |

MBF indicates myocardial blood flow. CVR indicates coronary vascular resistance. SD indicates standard deviation. Differences between preeclampsia and non-postpartum groups were calculated using unadjusted linear regression.

**Table S4. Association of time following delivery with myocardial flow reserve, myocardial blood flow, and coronary vascular resistance among postpartum individuals with preeclampsia (*n*=19).**

| | **Myocardial flow reserve** | | **Rest myocardial blood flow** | | **Stress myocardial blood flow** | | **Rest coronary vascular resistance** | | **Stress coronary vascular resistance** | |
|---|---|---|---|---|---|---|---|---|---|---|
| | *Beta (95% CI)* | *P-value* | *Beta (95% CI)* | *P-value* | *Beta (95% CI)* | *P-value* | *Beta (95% CI)* | *P-value* | *Beta (95% CI)* | *P-value* |
| No. of days postpartum at PET, per day | 0.05 (0.02 to 0.09) | 0.008 | -0.02 (-0.04 to -0.004) | 0.02 | 0.01 (-0.03 to 0.05) | 0.69 | 2.2 (-0.2 to 4.6) | 0.07 | -0.3 (-1.0 to 0.5) | 0.46 |

**Table S5. Subgroup analyses: Myocardial flow reserve, myocardial blood flow, coronary vascular resistance, and differences vs. non-postpartum individuals in preeclampsia with (*n*=8) or without (*n*=11) use of antihypertensive medication prior to imaging.** Women with preeclampsia who required use of antihypertensive medication on the morning of the study visit completed study assessments at a median [IQR] 14.5 [8.3, 16.0] days postpartum, and those who did not require antihypertensive medication completed assessments at 19.0 [16.0, 20.5] days postpartum (*P*=0.10).

| | **Preeclampsia with antihypertensive use on the morning of the imaging visit (*n*=8)** | | | **Preeclampsia without antihypertensive use on the morning of the imaging visit (*n*=11)** | | | **Non-post-partum (*n*=13)** |
|---|---|---|---|---|---|---|---|
| | Mean (SD) | Difference vs. non-postpartum | | Mean (SD) | Difference vs. non-postpartum | | Mean (SD) |
| | | *Beta (95% CI)* | *P-value* | | *Beta (95% CI)* | *P-value* | |
| **Myocardial flow reserve** | 2.65 (0.49) | -0.84 (-1.49 to -0.10) | 0.01 | 2.95 (0.82) | -0.55 (-1.22 to 0.12) | 0.10 | 3.49 (0.84) |
| **Rest MBF**, mL/min/g | 0.94 (0.20) | -0.02 (-0.18 to 0.14) | 0.81 | 0.94 (0.35) | -0.01 (-0.22 to 0.19) | 0.89 | 0.96 (0.15) |
| **Stress MBF**, mL/min/g | 2.49 (0.74) | -0.74 (-1.23 to -0.26) | 0.004 | 2.60 (0.59) | -0.64 (-1.04 to -0.24) | 0.003 | 3.24 (0.43) |
| **Rest CVR**, mmHg/mL/min/g | 101.5 (28.2) | 14.5 (-4.4 to 33.4) | 0.13 | 117.7 (46.5) | 30.7 (4.3 to 57.1) | 0.02 | 87.0 (16.0) |
| **Stress CVR**, mmHg/mL/min/g | 38.7 (11.6) | 12.9 (5.6 to 20.3) | 0.001 | 37.8 (10.7) | 12.0 (5.3 to 18.6) | 0.001 | 25.8 (6.2) |

MBF indicates myocardial blood flow. CVR indicates coronary vascular resistance. SD indicates standard deviation. Differences between preeclampsia and non-postpartum groups were calculated using unadjusted linear regression.

**Table S6. Association of clinical factors with PET indices among women with preeclampsia (*n*=19).**

| | Myocardial flow reserve | | Rest myocardial blood flow | | Stress myocardial blood flow | | Rest coronary vascular resistance | | Stress coronary vascular resistance | |
|---|---|---|---|---|---|---|---|---|---|---|
| | Beta (95% CI) | P-value | Beta (95% CI) | P-value | Beta (95% CI) | P-value | Beta (95% CI) | P-value | Beta (95% CI) | P-value |
| Age at PET, per year | 0.04 (-0.09 to 0.16) | 0.54 | 0 (-0.05 to 0.05) | 0.99 | 0.04 (-0.07 to 0.16) | 0.43 | 1.3 (-5.9 to 8.5) | 0.71 | -0.7 (-2.7 to 1.2) | 0.43 |
| BMI at PET, per $kg/m^2$ | -0.01 (-0.06 to 0.04) | 0.66 | -0.01 (-0.03 to 0.01) | 0.36 | -0.03 (-0.08 to 0.01) | 0.15 | 3.0 (0.6 to 5.5) | **0.02** | 0.9 (0.2 to 1.5) | **0.01** |
| BMI ≥30 $kg/m^2$ vs. <30 $kg/m^2$ at PET | -0.21 (-0.90 to 0.48) | 0.53 | -0.25 (-0.50 to 0.01) | 0.06 | -0.72 (-1.24 to -0.20) | **0.01** | 48.1 (17.0 to 79.2) | **0.004** | 14.9 (7.3 to 22.5) | **<0.001** |
| Gestational age at delivery, per week | 0.02 (-0.07 to 0.12) | 0.60 | 0.02 (-0.02 to 0.05) | 0.36 | 0.03 (-0.05 to 0.12) | 0.45 | 1.2 (-4.1 to 6.5) | 0.65 | -0.2 (-1.7 to 1.2) | 0.74 |
| Delivery <34 weeks vs. ≥34 weeks | -0.14 (-0.93 to 0.65) | 0.72 | -0.12 (-0.44 to 0.20) | 0.43 | -0.12 (-0.84 to 0.60) | 0.73 | -23.9 (-67.2 to 19.3) | 0.26 | 0 (-12.2 to 12.2) | 1 |
| Non-White vs. White race/ethnicity | -0.28 (-1.13 to 0.56) | 0.49 | 0.29 (-0.03 to 0.60) | 0.07 | 0.13 (-0.65 to 0.91) | 0.73 | 2.0 (-46.5 to 50.6) | 0.93 | -0.8 (-14.0 to 12.4) | 0.90 |
| Pre-pregnancy chronic hypertension vs. normotension | 0.05 (-0.80 to 0.91) | 0.90 | -0.06 (-0.41 to 0.29) | 0.72 | 0.09 (-0.69 to 0.87) | 0.82 | 7.9 (-40.4 to 56.3) | 0.73 | 2.3 (-10.8 to 15.5) | 0.71 |
| Primiparous (i.e., first pregnancy) vs. multiparous | -0.35 (-1.05 to 0.35) | 0.31 | 0.01 (-29 to 0.30) | 0.97 | -0.36 (-0.99 to 0.28) | 0.25 | -21.5 (-64.1 to 13.8) | 0.19 | 0.6 (-10.6 to 11.7) | 0.91 |
| Ever-smoking vs. never-smoking | -0.43 (-1.26 to 0.40) | 0.29 | -0.06 (-0.41 to 0.29) | 0.71 | -0.43 (-1.18 to 0.33) | 0.25 | 10.4 (-37.8 to 58.6) | 0.66 | 11.3 (-0.5 to 23.2) | 0.06 |

| | | | | | | | | | | |
|---|---|---|---|---|---|---|---|---|---|---|
| Use of aspirin in pregnancy vs. none | 0.43 (-0.29 to 1.15) | 0.23 | -0.10 (-0.40 to 0.20) | 0.49 | 0.24 (-0.44 to 0.91) | 0.47 | 28.0 (-12.1 to 68.1) | 0.16 | 2.7 (-8.8 to 14.2) | 0.63 |
| Max SBP before delivery, per mmHg | 0 (-0.02 to 0.02) | 0.71 | -0.01 (-0.01 to 0.00) | 0.13 | -0.02 (-0.03 to 0.00) | **0.04** | 0.5 (-0.6 to 1.5) | 0.39 | 0.2 (-0.1 to 0.5) | 0.16 |
| Max DBP recorded before delivery, per mmHg | -0.01 (-0.05 to 0.02) | 0.52 | 0 (-0.02 to 0.01) | 0.55 | -0.02 (-0.05 to 0.01) | 0.15 | 1.2 (-0.7 to 3.1) | 0.20 | 0.4 (-0.1 to 0.9) | 0.13 |
| HR with max SBP in pregnancy, per bpm | -0.01 (-0.03 to 0.01) | 0.32 | 0 (-0.01 to 0.01) | 0.49 | -0.01 (-0.03 to 0.00) | 0.12 | 0.6 (-0.6 to 1.8) | 0.32 | 0.3 (0.00 to 0.6) | 0.07 |
| Log(Maximum recorded proteinuria), per log(g) | 0 (-0.21 to 0.21) | 0.98 | -0.01 (-0.10 to 0.07) | 0.77 | -0.04 (-0.23 to 0.15) | 0.66 | -3.8 (-15.6 to 8.1) | 0.51 | -0.6 (-3.9 to 2.6) | 0.68 |
| Headache vs. none | -0.43 (-1.10 to 0.23) | 0.19 | -0.14 (-0.42 to 0.14) | 0.30 | -0.67 (-1.21 to -0.12) | **0.02** | 11.1 (-28.6 to 50.8) | 0.56 | 11.8 (2.8 to 20.9) | **0.01** |
| Vision changes vs. none | 0.52 (-0.29 to 1.33) | 0.19 | -0.13 (-0.47 to 0.21) | 0.43 | 0.28 (-0.49 to 1.05) | 0.45 | 19.0 (-28.6 to 66.6) | 0.41 | -2.7 (-15.8 to 10.4) | 0.67 |
| Pulmonary edema by CXR vs. none | 0.05 (-0.70 to 0.81) | 0.88 | -0.15 (-0.44 to 0.15) | 0.31 | -0.27 (-0.94 to 0.41) | 0.41 | 18.0 (-23.6 to 59.6) | 0.37 | 2.8 (-8.6 to 14.3) | 0.61 |
| ALT or AST >2x ULN vs. within normal limits | 0.12 (-0.59 to 0.82) | 0.73 | -0.05 (-0.34 to 0.24) | 0.73 | 0.11 (-0.53 to 0.76) | 0.72 | -18.1 (-57.1 to 21.0) | 0.34 | -3.1 (-13.9 to 7.7) | 0.55 |
| Minimum platelets <100K vs. ≥100K | -0.18 (-0.93 to 0.57) | 0.62 | 0.05 (-0.25 to 0.36) | 0.72 | -0.18 (-0.86 to 0.50) | 0.58 | -16.2 (-57.9 to 25.6) | 0.43 | -0.8 (-12.4 to 10.7) | 0.88 |
| Maximum recorded uric acid, per mg/dL | 0.06 (-0.23 to 0.35) | 0.66 | -0.01 (-0.12 to 0.11) | 0.92 | 0.03 (-0.25 to 0.28) | 0.90 | 2.0 (-14.4 to 18.4) | 0.80 | 0.0 (-4.7 to 4.7) | 1 |

| | | | | | | | | | | |
|---|---|---|---|---|---|---|---|---|---|---|
| Minimum hematocrit, per % | -0.02 (-0.11 to 0.07) | 0.59 | -0.01 (-0.05 to 0.02) | 0.44 | -0.02 (-0.11 to 0.06) | 0.53 | -1.1 (-6.1 to 4.0) | 0.66 | 0.7 (-0.6 to 2.0) | 0.27 |
| Newborn weight, per kg | 0.28 (-0.24 to 0.81) | 0.27 | 0.01 (-0.21 to 0.24) | 0.89 | 0.13 (-0.36 to 0.63) | 0.58 | 13.4 (-16.8 to 43.5) | 0.36 | -2.1 (-10.4 to 6.2) | 0.60 |

**Table S7. Characteristics of the study cohort with echocardiographic data.**

| | Preeclampsia, postpartum (*n*=17) | Normotensive, postpartum (*n*=7) | P-value |
|---|---|---|---|
| Age at study visit | 32.6 (2.7) | 33.0 (4.0) | 0.82 |
| Race/ethnicity<br>• Asian<br>• Black<br>• Hispanic<br>• White | <br>0 (0%)<br>2 (11.8%)<br>2 (11.8%)<br>13 (76.5%) | <br>2 (28.6%)<br>0 (0%)<br>1 (14.3%)<br>4 (57.1%) | 0.17 |
| Chronic hypertension | 4 (23.5%) | 0 (0%) | 0.28 |
| Chronic diabetes mellitus | 0 (0%) | 0 (0%) | 1 |
| Former smoking history<br>• Within past 12 months | 4 (23.5%)<br>1 (5.9%) | 0 (0%)<br>0 (0%) | 0.28<br>1 |
| History of hypertensive disorder in previous pregnancy | 3 (17.6%) | 0 (0%) | 0.53 |
| Parity<br>• 1<br>• 2<br>• 3+ | <br>11 (64.7%)<br>3 (17.6%)<br>3 (17.6%) | <br>3 (42.9%)<br>2 (28.6%)<br>2 (28.6%) | 0.57 |
| Pre-pregnancy body mass index | 27.5 [22.7, 32.3] | 24.0 [20.8, 25.8] | 0.10 |
| Gestational diabetes in index pregnancy | 1 (5.9%) | 0 (0%) | 1 |
| Use of aspirin for preeclampsia prevention in index pregnancy | 6 (35.3%) | 0 (0%) | 0.13 |
| Gestational age at delivery, weeks + days | 31+2<br>[29+4, 35+4] | 39+0<br>[38+5.75, 39+6.25] | <0.001 |
| Mechanism of delivery<br>• Spontaneous vaginal<br>• Assisted vaginal<br>• Cesarean section | <br>1 (5.9%)<br>0 (0%)<br>16 (94.1%) | <br>5 (71.4%)<br>1 (14.3%)<br>1 (14.3%) | <0.001 |
| Days postpartum at study visit | 15.5 (7.9) | 13.7 (7.8) | 0.63 |
| Body mass index at study visit | 28.9<br>[26.3, 35.8] | 22.7<br>[22.6, 30.9] | 0.08 |
| Rest systolic blood pressure at study visit | 138.2 (20.0) | 104.4 (14.3) | 0.002 |
| Rest diastolic blood pressure at study visit | 77.9 (12.6) | 62.2 (9.4) | 0.002 |
| Rest heart rate at study visit | 72.5 (11.8) | 55.6 (5.9) | 0.001 |

**Table S8. Correlation of myocardial flow reserve, myocardial blood flow, and coronary vascular resistance with echocardiographic parameter among postpartum individuals with preeclampsia (*n*=19).**

| | **Myocardial flow reserve** | | **Rest MBF** | | **Stress MBF** | |
|---|---|---|---|---|---|---|
| | *r* | P-value | *r* | P-value | *r* | P-value |
| Septal wall, mm | -0.35 | 0.16 | 0.18 | 0.50 | -0.26 | 0.32 |
| Posterior wall, mm | -0.34 | 0.18 | -0.03 | 0.91 | -0.49 | **0.04** |
| Relative wall thickness | -0.33 | 0.20 | 0.05 | 0.86 | -0.39 | 0.13 |
| LVEDD, mm | 0.11 | 0.69 | -0.21 | 0.41 | -0.13 | 0.62 |
| LVESD, mm | 0.41 | 0.10 | -0.26 | 0.32 | 0.19 | 0.45 |
| LV ejection fraction, % | -0.37 | 0.14 | 0.58 | **0.02** | 0.21 | 0.42 |
| LA volume, mL | -0.27 | 0.30 | -0.26 | 0.32 | -0.56 | **0.02** |
| Mitral E, cm/s | -0.35 | 0.17 | 0.22 | 0.40 | -0.24 | 0.36 |
| Mitral A, cm/s | -0.32 | 0.22 | 0.32 | 0.22 | -0.23 | 0.37 |
| Mitral E/A ratio | -0.05 | 0.85 | 0.00 | 0.99 | 0.08 | 0.77 |
| Lateral e’, cm/s | -0.16 | 0.54 | 0.53 | **0.03** | 0.30 | 0.24 |
| Septal e’, cm/s | -0.24 | 0.35 | 0.37 | 0.14 | 0.11 | 0.69 |
| Average E/e’ ratio | -0.04 | 0.88 | -0.38 | 0.14 | -0.45 | 0.07 |
| TR velocity, cm/s | 0.37 | 0.21 | 0.03 | 0.93 | 0.30 | 0.33 |
| S’, cm/s | -0.27 | 0.30 | 0.34 | 0.18 | 0.05 | 0.86 |
| TAPSE, cm | -0.09 | 0.72 | 0.30 | 0.25 | 0.12 | 0.65 |
| RV fractional age change, % | 0.08 | 0.76 | -0.02 | 0.95 | 0.15 | 0.57 |
| Basal LV longitudinal strain, % | -0.13 | 0.61 | 0.23 | 0.37 | -0.03 | 0.91 |
| Mid-LV longitudinal strain, % | -0.09 | 0.74 | 0.13 | 0.61 | -0.01 | 0.96 |
| Apical longitudinal strain, % | 0.10 | 0.70 | 0.02 | 0.95 | 0.00 | 0.99 |
| Global longitudinal strain, % | 0.07 | 0.79 | 0.01 | 0.97 | 0.01 | 0.96 |

**Table S9. Correlation of angiogenic biomarkers with PET indices.**

| | Myocardial flow reserve | | Rest myocardial blood flow | | Stress myocardial blood flow | | Rest coronary vascular resistance | | Stress coronary vascular resistance | |
|---|---|---|---|---|---|---|---|---|---|---|
| | *r* | P-value | *r* | P-value | *r* | P-value | *r* | P-value | *r* | P-value |
| Log(sFlt-1/PlGF ratio) | -0.45 | 0.05 | 0.71 | <0.001 | 0.22 | 0.36 | -0.55 | 0.01 | -0.16 | 0.51 |
| Log(sFlt-1) | -0.46 | 0.06 | 0.56 | 0.02 | 0.05 | 0.82 | -0.45 | 0.13 | -0.01 | 0.91 |
| Log(PlGF) | -0.34 | 0.15 | 0.23 | 0.35 | -0.15 | 0.55 | -0.06 | 0.81 | 0.25 | 0.31 |
